# Atypical influence of biomechanical knowledge in Complex Regional Pain Syndrome-Towards a different perspective on body representation

**DOI:** 10.1101/2021.10.05.21264512

**Authors:** L. Filbrich, C. Verfaille, G. Vannuscorps, A. Berquin, O. Barbier, X. Libouton, V. Fraselle, D. Mouraux, V. Legrain

## Abstract

Part of the multifaceted pathophysiology of Complex Regional Pain syndrome (CRPS) has been ascribed to a lateralized maladaptive neuroplasticity in sensorimotor cortices, a finding that has been corroborated by behavioral studies indicating that CRPS patients indeed present difficulties in mentally representing their painful limb. Hand laterality judgment tasks (HLT) are widely used to measure such difficulties, with the laterality of hand stimuli corresponding to the affected hand judged more slowly than the one of hand stimuli corresponding to the unaffected hand. Importantly, the HLT is also regularly used in the rehabilitation of CRPS and other chronic pain disorders, with the aim to activate motor imagery and, consequently, restoring the cortical representation of the limb. The potential of these tasks to elicit motor imagery is thus critical to their use in therapy. Yet, the influence of the biomechanical constraints (BMC) on HLT reaction time, supposed to reflect the activation of motor imagery, is rarely verified. In the present study we investigated the influence of the BMC on the perception of hand postures and movements. The results of a first experiment, in which a HLT was used, showed that CRPS patients were significantly slower than controls in judging hand stimuli, whether or not the depicted hand corresponded to their affected hand, but that their performance did not differ from controls when they judged non-body stimuli. Results regarding reaction time patterns reflecting the BMC were inconclusive in CRPS and controls, questioning the validity of the task in activating motor imagery processes. In a second experiment we therefore directly investigated the influence of implicit knowledge of upper-limb BMC on perceptual judgments of hand movements with the apparent body movement perception task. Participants judge the perceived path of movement between two depicted hand positions, with only one of the two proposed paths that is biomechanically plausible. While the controls chose the biomechanically plausible path most of the time, CRPS patients did not, indicating that the perception and/or use of the BMC seems to be disturbed in CRPS. These findings show a non-lateralized body representation impairment in CRPS, which might be related to difficulties in using correct knowledge of the body’s biomechanics. Most importantly however, our results, in agreement with previous studies, indicate that it seems highly challenging to measure motor imagery and the indexes of BMC with the classical HLT task, which has important implications for the rehabilitation of chronic pain with these tasks.

## 1. Introduction

Patients suffering from Complex Regional Pain Syndrome (CRPS) are characterized by severe and continuous pain in one limb, which often develops after minor or moderate trauma and which is disproportionate to the triggering event (Marinus *et al*., 2011). Sensory symptoms are accompanied by various autonomic, trophic and motor symptoms (Harden *et al*., 2010). CRPS is difficult to treat, since its pathophysiology is complex, involving different mechanisms, at different stages and individual time frames, such as neurogenic inflammation, vasomotor dysfunction as well as structural and functional changes at the cortical level (Marinus *et al*., 2011; Birklein and Schlereth, 2015; Birklein and Dimova, 2017). These cortical changes have been mainly interpreted as reflecting a *maladaptive neuroplasticity* of the cortical representation of the affected limb, mostly in the primary and secondary somatosensory (SI, SII) as well as primary motor (MI) cortices (e.g. Juottonen *et al*., 2002; Maihofner *et al*., 2003, 2004; Pleger *et al*., 2004; Pleger *et al*., 2005; Krause *et al*., 2006; Pleger *et al*., 2006; Maihofner *et al*., 2007; Pleger *et al*., 2014; Pfannmoller *et al*., 2019; however, see Mancini *et al*., 2019 for contrasting results;). Consequently, cognitive difficulties of CRPS patients in mentally representing and perceiving their affected limb have been extensively investigated as indexing this cortical pathophysiology of CRPS and are now recognized as a typical feature of the CRPS symptomatology (Reinersmann *et al*., 2013). Those studies have also driven the development of rehabilitation techniques promoting the restoration of cortical sensorimotor representations of the affected limb in CRPS (Moseley and Flor, 2012). Such problems in body representation include feelings of disownership over the affected limb as well as distortions in representing its size, shape and position (Halicka *et al*., 2020).

One specific task that is regularly used to investigate body representation difficulties in CRPS is the hand laterality task (HLT; Cooper and Shepard, 1975; Parsons, 1987). In this task, images of hands are presented and participants have to judge as quickly and accurately as possible whether they see a left or a right hand. The different hand stimuli are presented in various rotation directions, i.e. towards or away from the body midline, and orientations, i.e. with different degrees of angular deviation from a canonical hand stimulus at 0°. When tested with the HLT, upper-limb CRPS patients have been shown to be much slower in judging the laterality of hand stimuli corresponding to their affected upper limb than of hand stimuli corresponding to their unaffected upper limb (Moseley, 2004b; Reid *et al*., 2016), while such a difference between left and right hands was not observed for healthy control participants (Moseley, 2004b). This observation seems to support the idea that CRPS patients have difficulties in generating and manipulating a cortical representation of their affected limb. However, the results of other studies that used the HLT to test body representation in CRPS indicate that this effect might not be as consistently found as previously thought. Indeed, some studies show that CRPS patients are only slower in recognizing the laterality of a depicted hand corresponding to the affected limb when its orientation deviates the most from its canonical presentation (i.e. 180°) (Schwoebel *et al*., 2001; Schwoebel *et al*., 2002). Other studies found increased reaction times (RTs) for both hands as compared to controls (Reinersmann *et al*., 2010; Bultitude *et al*., 2017; Wittayer *et al*., 2018) or no difficulties in judging hand laterality at all in CRPS (Reinersmann *et al*., 2012; Breimhorst *et al*., 2018).

The HLT is hypothesized to more specifically test the body schema, an online sensorimotor representation of the body. This unconscious and dynamic representation of the relative position of the body parts would interact with the motor systems to generate and guide actions (Schwoebel and Coslett, 2005). The body schema would thus underlie real movement, but it has been hypothesized that this would also be the case for imagined movements, as in the HLT (Parsons, 1994; Schwoebel *et al*., 2004; Schwoebel and Coslett, 2005). Indeed, it has been shown that the HLT can prime motor imagery, i.e., in this case, the mental manipulation of body parts and simulation of hand movement from a first-person perspective. Consequently, the RT to judge the laterality of the depicted hand is considered as the time that is necessary to perform a mental rotation of one’s own hand towards the depicted position of the stimulus hand. This has been based on the observation that participants’ RTs on the HLT depend on the current position of their own hand (e.g. Parsons, 1994; Sirigu and Duhamel, 2001; Ionta *et al*., 2007), as well as on the human body’s biomechanical constraints on movements, i.e. the laws of mechanics that are applied to the range of motion of the different body parts. Specifically, in the HLT, when the movement to mentally rotate the physical hand to the position of the hand stimulus is close to the biomechanical constraints of the hand, the RT increases. For example, images of hands in a laterally-rotated position (i.e. rotated away from the mid-sagittal plane of the body) are generally judged slower than those in medially-rotated position (i.e. rotated toward the mid-sagittal plane), an effect called the Medial-Over-Lateral-Advantage (MOLA effect, see Funk and Brugger, 2008; Vannuscorps *et al*., 2012). Indeed, a medial rotation with the real hand would often be less constrained, and therefore take less time, than a lateral rotation (Parsons, 1987, 1994).

The presence of such biomechanical indexes reflected in the RTs at the HLT has thus generally been taken as evidence that motor imagery processes have been activated during the task (although see Vannuscorps *et al*., 2012). However, in the aforementioned HLT studies in CRPS, these biomechanical indexes have not been systematically analysed. One reason for inconsistent results between these studies could thus possibly be that not all of them trigger motor imagery processes with their task. It is indeed known that under some circumstances, participants can switch to alternative strategies, such as visual imagery, another form of mental imagery, during the HLT (e.g. Kosslyn *et al*., 1998; Daprati *et al*., 2010; ter Horst *et al*., 2010). Visual imagery is commonly used in tasks in which objects, such as figures, numbers or letters, have to be mentally rotated (e.g. Shepard and Metzler, 1971). In the case of visual imagery, the depicted hands would not be treated as one’s body part, but rather as an external object whose orientation in space could be evoked by a movement from a third person perspective or without simulating an actual motor action (Sirigu and Duhamel, 2001). Consequently, the above-described biomechanical indexes in RTs should not be observed if visual imagery is used to solve the HLT (e.g. de Lange *et al*., 2005).

The aim of the present study was to investigate hand representation in CRPS with two experiments, by focusing on the influence of the biomechanical constraints of the hands on the perception of hand postures and movements. In a first experiment, CRPS patients and matched controls performed a HLT on left and right hand stimuli. Based on the literature, we hypothesized that the CRPS patients would be slower in judging hand stimuli corresponding to their affected hand as compared to their unaffected hand. Furthermore, we also investigated whether, and in which conditions, both CRPS and control participants’ judgments were influenced by the hand biomechanics when performing the laterality judgments, by testing the presence of the MOLA effect on the RT. Importantly, all participants performed the task also on control stimuli, i.e. pictures of letters and houses, which are, as opposed to the hand stimuli, supposed to induce visual imagery processes instead of motor imagery. Consequently, we expected to see these effects only for hand stimuli. Finally, all the participants performed the task according to two different task instructions, either inducing motor or visual imagery processes. In a second experiment we investigated the influence of patients’ and control participants’ knowledge of the biomechanical constraints of the upper limbs on their perception of hand movements, with a test based on the apparent motion paradigm (e.g. Shiffrar and Freyd, 1990; Chatterjee *et al*., 1996). In this task participants have to select the perceived path of movement between two depicted hand positions, of which only one of the two proposed paths is biomechanically plausible. We hypothesized that if CRPS patients have a distorted representation, perception and/or use of the biomechanics of their affected upper limb, they should be different from control participants in their probability of choosing the biomechanically plausible vs. implausible path of movement, especially when the depicted limb corresponds to their affected limb.

## 2. Methods

### 2.1 Participants

Nineteen patients with upper-limb CRPS participated in the study. Data from one patient were excluded from the analyses because of incomplete testing due to excessive fatigue. Sixteen patients participated in Experiment 1 and 12 in Experiment 2. Patient characteristics are presented in Table 1. Sixteen healthy volunteers participated as control participants in Experiment 1 (14 women, 53±6.9 years old, range: 35-64 years, all right-handed) and thirteen in Experiment 2 (8 women, 52±9.7 years old, range: 28-64 years, all right-handed). For all participants in both experiments, exclusion criteria were the presence of any neurological and severe psychiatric disorder, any unresolved orthopedic injuries as well as uncorrected vision difficulties. Additionally, for control participants, exclusion criteria included the presence of chronic pain and upper-limb trauma during the past year. Three of the patients had previously followed a graded motor imagery training (Moseley *et al*., 2012) of at least two weeks in the context of their physiotherapy sessions and at home (participants 05, 12 and 13 in Table 1). Experimental procedures were approved by the local ethics committee (EudraCT: B403201214265) and conform to the Declaration of Helsinki. All patients provided written informed consent and received a financial compensation for their participation.

**Table 1.**
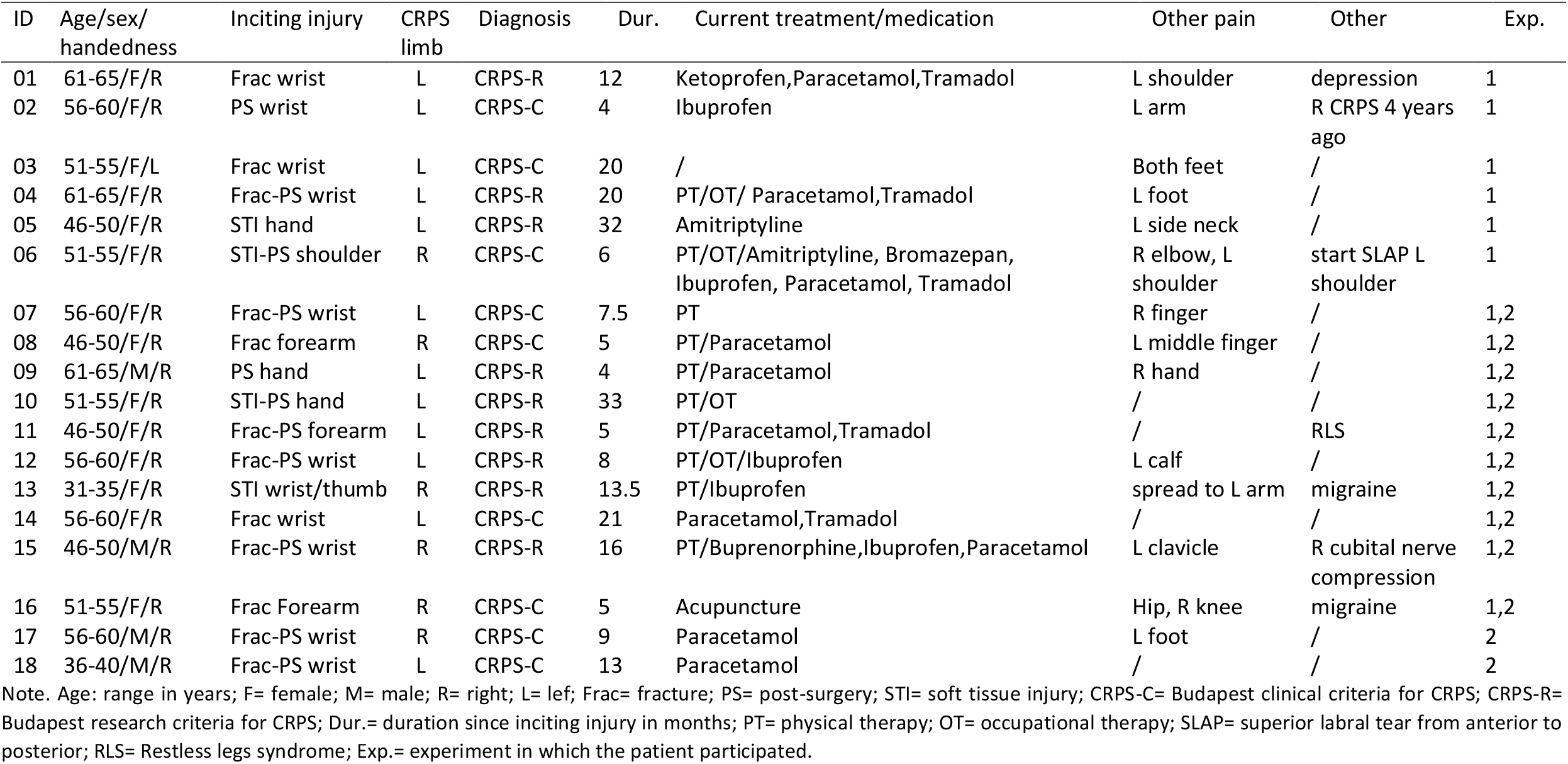
Patient characteristics

### 2.2 Experiment 1: Mental imagery task

#### 2.2.1 Stimuli and apparatus

Visual stimuli were pictures of open hands in back-view and images of a house with a chimney or of the letter L (Fig. 1). Stimuli were presented on a grey background (RGB: 128,128,128) using E-prime 2.0 (Psychology Software Tools, Inc., Pittsburgh, PA, USA) on a 16.6” LCD screen (1280*1024 resolution, 75Hz refresh rate, anti-glare filter). Images were built and resized with Photoshop CC 2015, appearing on the screen with a stimulus size of 13 cm height and 8.5 cm width. Pictures and images could be horizontally flipped so that hands appeared as either left or right hands, houses appeared with a chimney to either the left or right side, and letters L with the horizontal bar placed either at the left or right side of the vertical bar (*laterality* conditions). Pictures and images were presented with different *orientation* angles (0°, 45°, 90°, 135° or 180° in the picture plane) and with *rotations* in two possible directions (clockwise or anticlockwise from 0° to 180°, recoded into medial and lateral rotations, see Fig. 1). In total, for each of the 3 types of stimuli, 20 different stimuli were created (2 lateralities x 5 orientations x 2 rotations).

**Figure 1.**
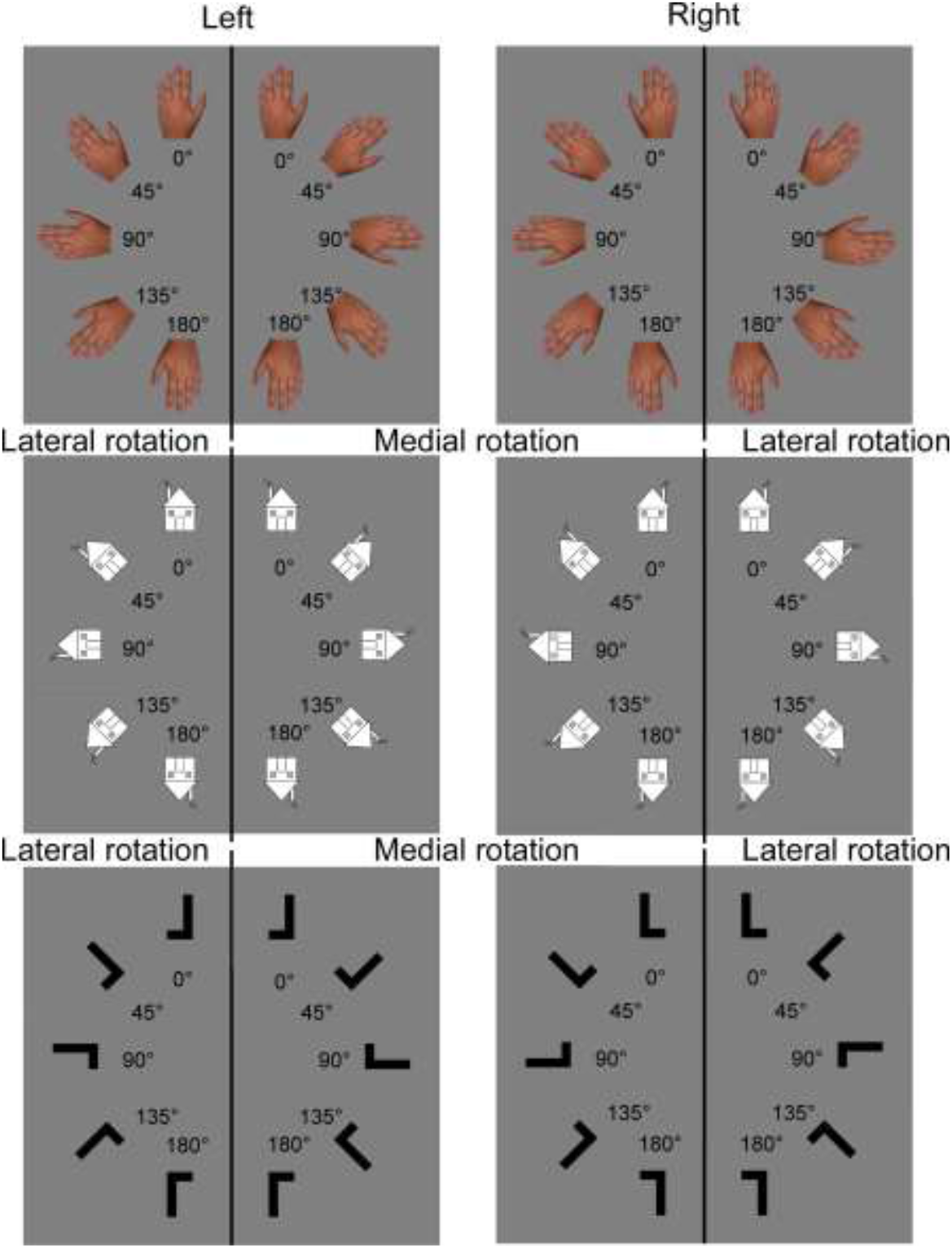
Stimuli of the mental imagery task. Stimuli result from the combination of stimulus type (hand, house or letter), laterality (left or right), rotation (medial or lateral) and orientation (0°, 45°, 90°, 135° or 180°). For pictures of hands, clockwise rotations of the left hand and anticlockwise rotations of the right hand correspond to medial rotations. Anticlockwise rotations of the left hand and clockwise rotations of the right hand correspond to lateral rotations. For the purpose of statistical analyses, rotation conditions of the house and letter images were relabeled in order to match conditions and the response codes of the hand pictures. Accordingly, for images of houses with a right-sided chimney and the L with the horizontal bar at the right, clockwise rotations are considered as lateral rotations and anticlockwise rotations as medial rotations. The reverse is the case for images of houses with a left-sided chimney and the L with the horizontal bar at the left.

#### 2.2.2 Procedure

Participants sat approximatively 60 cm from a computer screen with their head on a chinrest. CRPS patients used their unaffected hand to respond on an azerty keyboard placed at 31 cm from the edge of the table, while the affected one rested passively palm down on their tight. Similarly, each control participant performed the task with the same hand as the one used by the CRPS patient to whom he/she was individually matched. The responding hand and the keyboard were hidden from sight by a black cloth attached to the chinrest.

Participants performed 12 blocks of stimuli, divided into two sessions of six blocks. Each block consisted of eight randomly presented repetitions of the 20 stimuli of a single stimulus type and each session comprised two blocks of each stimulus type. During one session, the two blocks of the same stimulus type were presented directly one after the other, but the order of presentation of the different stimulus types was randomized. A trial started with the presentation of a black fixation cross. After 500 ms, the fixation cross was replaced by a stimulus that stayed on screen until the participant responded by pressing one of two keys on the keyboard, which initiated the next trial. During one session, participants were asked to judge the spatial laterality of the stimuli (*laterality judgment* task). More precisely, they judged whether the hand stimulus corresponded to a left or a right hand, whether the chimney was on the left or the right side of the house, or whether the horizontal bar of the letter L was on the left or the right side of the vertical bar (*b* key = left, *n* key = right, for all stimuli). In the other session, participants were asked to match each of the presented stimuli to an initial probe stimulus (*matching* task). Each block was preceded by the presentation of a right probe stimulus in the 0° orientation angle and participants judged whether each following stimulus of the same type was the same or not as the probe stimulus in terms of laterality (*b* key = same, *n* key = different). Participants were instructed to respond both as quickly and as accurately as possible. The order of the sessions was counterbalanced. For each session, before and after each pair of blocks of a same stimulus type, CRPS patients rated the pain in their affected limb on a numeric rating scale ranging from 0-10 (with 0= no pain and 10= worst pain imaginable).

In each session, before changing the type of stimulus, participants performed a training block on all 20 stimuli of one type. In order to pass to the first experimental block of the corresponding stimulus type, participants were required to accurately judge at least 16 out of the 20 presented stimuli. If this performance was not achieved, the training was repeated. Duration of one experimental block was two to five minutes.

#### 2.2.3 Measures

Performance was measured by means of reaction time (RT, in ms) and by accuracy, i.e., the percentage of correct responses (relative to the total of stimuli for each condition). Only trials with correct responses were included in RT measures. Trials with RTs lower than 300 ms and higher than 10 000 ms were removed from analyses (<1% removed in total, no aberrant data remained). Before the analyses, stimuli corresponding to pictures of hands were recoded according to their correspondence to the affected (i.e. ipsilateral) vs. the unaffected limb (i.e. contralateral) for each CRPS patient. To match condition labelling and facilitate statistical comparisons, house and letter images were similarly recoded for CRPS patients, and for control participants all stimuli were recoded according to the affected side of the CRPS patient to whom they were individually matched.

#### 2.2.4 Data analysis

ANOVAs for repeated measures were performed on the accuracy and RT data with *stimulus type* (hand vs. house vs. letter), *laterality* (ipsilateral vs. contralateral), *rotation* (medial vs. lateral), *orientation* (0° vs. 45° vs. 90° vs. 135° vs. 180°) and *task* (laterality judgment vs. matching) as within-participant factors and *group (CRPS vs. control)* as between-participant factor. Effect size was measured using partial Eta squared. Greenhouse-Geisser corrections and contrast analyses were performed when needed and the significance level was set at p ≤ .05.

Additional analyses regarding pain ratings during the task are described in the Supplementary materials.

### 2.3 Experiment 2: Apparent body movement perception task

#### 2.3.1 Stimuli and apparatus

Visual stimuli are from the study of Vannuscorps and Caramazza (2016) and were presented using E-Prime 2.0 on the same screen as in Experiment 1. Stimuli consisted of four pairs of pictures depicting an actor with his right (2 pairs) or left (2 pairs) upper limb raised at 90° relative to the trunk. The first picture of the pair depicts the forearm in supine position and the wrist either in flexion or in extension. The second picture depicts the pronation of the forearm by a medial rotation movement. The sequential presentation of the two pictures of a pair gives the illusion of movement of the forearm and the hand between the two positions, a movement that can be visually perceived according to two possible rotation paths: a long path (270°), illustrating the medial rotation that is biomechanically possible to execute, and a short path (90°), illustrating a lateral rotation that is biomechanically impossible (Nordin and Frankel, 2001). The stimuli can be visualized at http://www.testable.org/experiment/7/919310/start.

#### 2.3.2 Procedure

The experimental set-up was similar to Experiment 1. Before the task, participants were asked to report what they saw in a duck-rabbit illusion (Brugger and Brugger, 1993), to illustrate that the perception of a same stimulus could differ from person to person and time to time. They were told that, as the duck-rabbit illusion, the following experiment was an example of such perceptual ambiguity and that there was no correct or incorrect response. The use of this illusion aimed at preventing participants from questioning the plausibility of the perceived movement paths during the experiment (see Vannuscorps and Caramazza, 2016 for a detailed description).

During the experiment, participants were presented with 4 blocks of 40 trials each. Each trial started with a blank screen for 500 ms followed by a sequence of four successive presentations of the same pair of pictures. The presentation of the two pictures of the pair were separated by a blank screen. Five presentation speeds were used: the respective durations of presentation of the pictures and the blank screen were 100ms-50ms (speed 1), 150ms-100ms (speed 2), 200ms-150ms (speed 3), 250ms-200ms (speed 4) and 300ms-250ms (speed 5). A blank screen appeared after the last picture of the sequence, which was then followed after 1s by a figure presenting the actor’s two hand positions shown during the trial and two possible paths (i.e. short vs. long) of apparent movement between these two hand positions. The two movement paths were labelled ‘A’ or ‘B’, respectively, the assignment of one letter to one particular path being counterbalanced across trials. Participants were asked to choose the path corresponding to the movement they perceived during the sequential presentation of the two pictures. This response stayed on screen until the participants responded by pressing one of two keys on the keyboard corresponding to the two response labels. Once the response was provided, the next trial started (see http://www.testable.org/experiment/7/919310/start). Each trial was composed of the combination of three different variables: *laterality* of the upper limb (left vs. right), *position* of the hand (flexed vs. extended wrist) and *speed* (1 to 5). Each of these 20 possible combinations was repeated twice per block, i.e. eight times in total, and were randomly presented. Before the experiment, participants completed a training of five trials. One block lasted approximately 5 minutes.

#### 2.3.3 Measures

For each laterality and speed combination, we measured the percentage of trials in which the long path was chosen, i.e. in which the participants chose the biomechanically plausible path as the perceived movement. Data from the two positions of the hand were averaged. Regarding the laterality factor, CRPS patients’ data were recoded to match the actor’s raised upper limb with their affected or unaffected upper limb. For control participants, laterality was coded according to the CRPS patient to whom they were matched. For example, a stimulus showing the actor raising his left arm was coded as “affected” for a patient with left-sided CRPS and his/her matched control.

#### 2.3.4 Data analysis

To test whether the percentage of long, i.e. biomechanically plausible, paths chosen would be different between the two groups and whether this would be modulated by the laterality of the depicted upper limb, an ANOVA with *speed* (1 to 5) and *laterality* (affected vs. unaffected) as within-participant factors and *group* (CRPS vs. control) as between-participant factor was performed. To test whether participants perceived one of the two paths significantly more often than the other path, the percentage of trials in which the long path was chosen was compared to the percentage of trials in which the short path was chosen by means of paired-samples t-tests. Effect sizes were measured using Cohen’s d for t-tests and partial Eta squared for ANOVAs. Contrast analyses and Greenhouse-Geisser corrections of degrees of freedom were performed when necessary. The significance level was set at p ≤ .05.

## 3. Results

### 3.1 Experiment 1: Mental imagery task

All the main effects and interactions that reached significance in the RT ANOVA can be reviewed in Table 2, whereas the complete results of the ANOVAs can be found in the Supplementary materials Table S1 and Table S2. Because of the multitude of factors, only the results regarding our specific hypotheses for RTs are specified here. Results not directly related to our RT hypotheses, the accuracy data or pain ratings can be found in the Supplementary materials.

**Table 2.** Significant results of the ANOVAs on the RT data with stimulus type (hand vs. house vs. letter), laterality (ipsilateral vs. contralateral), rotation (medial vs. lateral), orientation (0° vs. 45° vs. 90° vs. 135° vs. 180°) and task (laterality judgment vs. matching) as within-participant factors, and group (CRPS vs. control) as between-participant factor

| Reaction time |  |  |  |  |
| --- | --- | --- | --- | --- |
| **Factors** | **F** | **df** | **p** | **$\eta^2_p$** |
| Group | 7.95 | 1, 30 | .008 | .21 |
| Stimulus | 63.58 | 1.26, 37.95 | .000 | .68 |
| Orientation | 197.73 | 1.43, 42.86 | .000 | .87 |
| Group x Stimulus | 12.28 | 1.26, 37.95 | .001 | .29 |
| Group x Rotation | 8.38 | 1, 30 | .007 | .22 |
| Group x Rotation x Orientation | 3.51 | 2.33, 70.06 | .029 | .10 |
| Group x Rotation x Laterality | 5.84 | 1, 30 | .022 | .16 |
| Stimulus x Orientation | 8.60 | 3.05, 91.47 | .000 | .22 |
| Stimulus x Orientation x Rotation | 3.80 | 4.73, 141.80 | .004 | .11 |
| Stimulus x Task | 4.49 | 1.48, 44.48 | .026 | .13 |

Note. df= degrees of freedom.

We first tested whether CRPS patients would be slower than control participants when judging the laterality of hand stimuli, and whether this would be specifically the case when judging ipsilateral hand stimuli, i.e. hand stimuli corresponding to their affected upper limb. The ANOVA indeed revealed an interaction between *group* and *stimulus type* (F(1.26,37.95) = 12.28, *p* = .001, η^2^_P_ = .29), which was however not modulated by the laterality of the depicted hand, since the interaction between *group, laterality* and *stimulus type* was not significant (F(1.63,48.97) = .58, *p* = .530, η^2^_P_ = .02). Contrast analyses showed that this *group x stimulus type* interaction could be related to the fact that the CRPS patients were significantly slower in responding to hand stimuli than the control participants (mean ± sd, CRPS: 1955±521 ms; Controls: 1298±301 ms; F(1,30)= 11.94, *p* = .002, η^2^_P_ = .29) and that such a difference between the groups was not significant for house and letter stimuli (all F ≤ 1.90, *p* ≥ .179, η^2^_P_ ≤ .06; Fig. 2). Contrasts regarding differences between the types of stimuli in each of the groups are detailed in Fig. 2 and in the Supplementary material.

**Figure 2.**
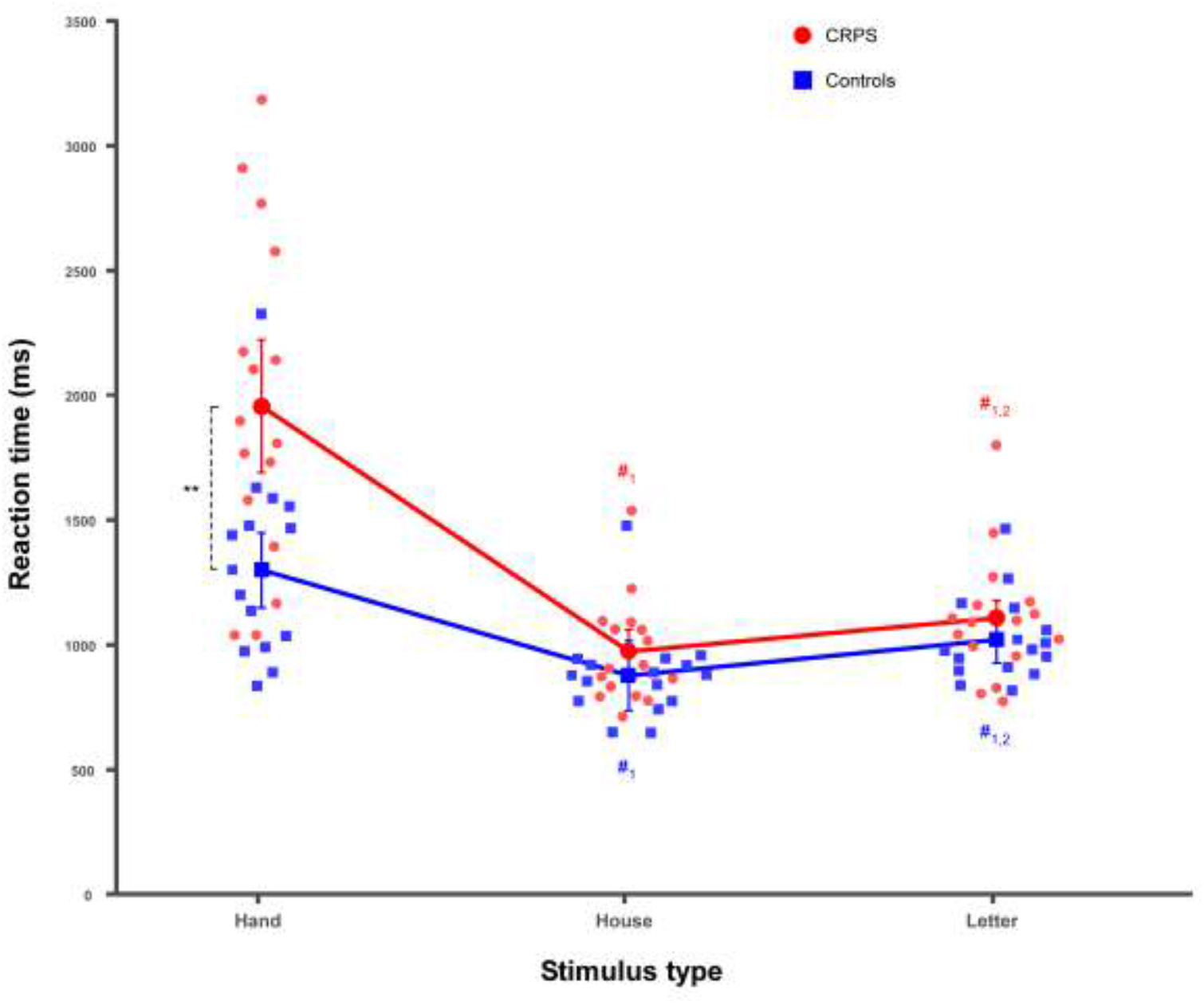
Mean reaction times (RT) according to group and stimulus type. CRPS patients are represented in red circles and control participants in blue squares. Smaller and lighter colored circles/squares represent the individual data. Error bars represent the 95% confidence intervals adapted according to the method of Cousineau (Cousineau, 2005). **p ≤ .01. #1 = significantly different from hand stimuli, #2 = significantly different from house stimuli.

Additionally, we tested for the presence of a specific RT pattern indexing the biomechanical constraints of the hands, i.e. the MOLA effect (RT medial stimuli ≠ lateral stimuli). The results showed that the *stimulus type* and *rotation* factors interacted with the *orientation* factor (F(4.73,141.79) = 3.80, *p* = .004, η^2^_P_ = .11) (Fig. 3). Contrast analyses revealed that only for stimuli presented at 45° and 135° the RTs were modulated by an interaction between stimulus type and rotation (**45°**: F(1.24,37.29) = 4.63, *p* = .03, η^2^_P_ = .13; **135°**: F(1.63,48.74) = 4.68, *p* = .019, η^2^_P_ = .14; all other F ≤ 2.88, *p* ≥ .076, η^2^_P_ ≤ .09). Indeed, at 45°, only hand stimuli were significantly judged faster when they were medially rotated than when they were laterally rotated (F(1,30) = 6.28, *p* = .018, η^2^_P_ = .17; all other F ≤ 3.38, *p* ≥ .076, η^2^_P_ ≤ .10). Surprisingly, when stimuli were turned at 135°, medially rotated stimuli were only judged faster than laterally rotated stimuli when house stimuli were presented (F(1,30) = 12.28, *p* = .001, η^2^_P_ = .29 ; all other F ≤ 1.87, *p* ≥ .181, η^2^_P_ ≤ .06).

**Figure 3.**
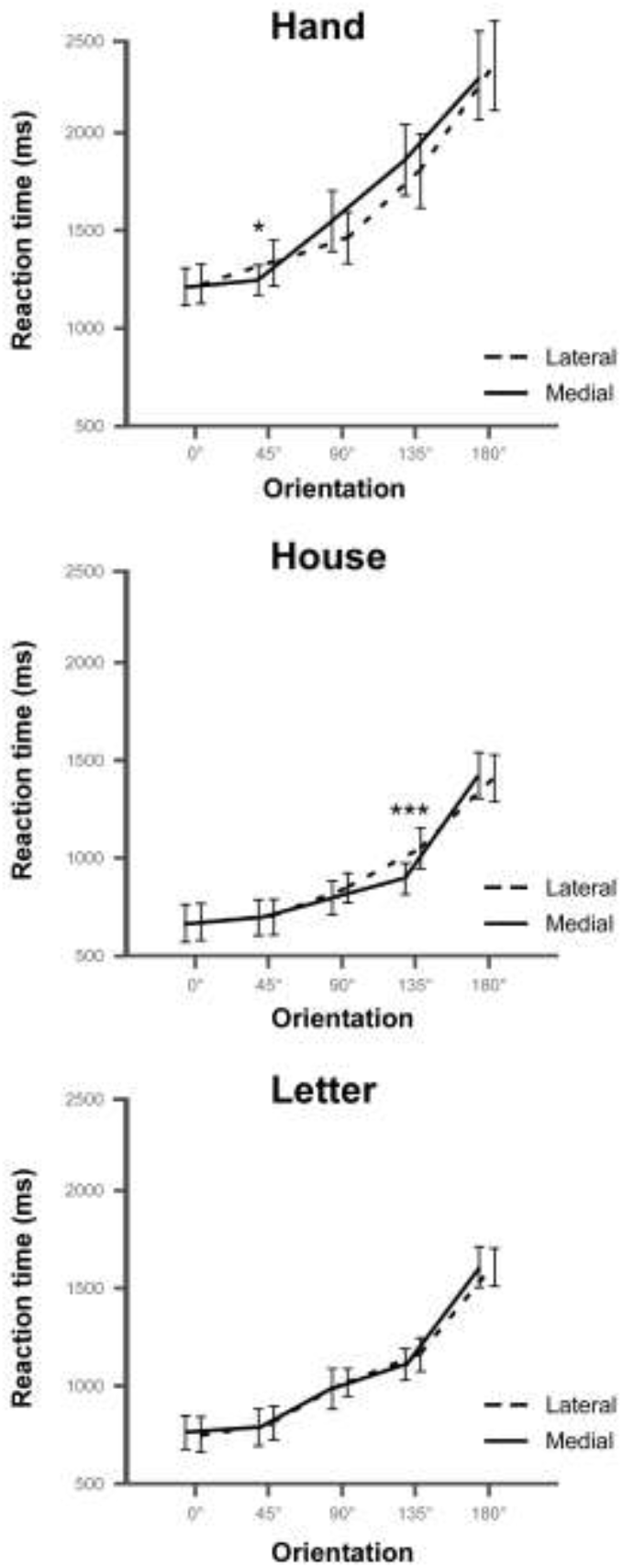
Mean reaction times (RT) according to stimulus type, rotation and orientation of the stimuli. The lines represent the comparison of RTs between the lateral (dashed lines) and medial (plain lines) rotations according to the orientation of the stimuli (i.e. 0° vs. 45° vs. 90° vs. 135° vs. 180°) for each type of stimulus (i.e. hand vs. house vs. letter). Error bars represent the 95% confidence intervals adapted according to the method of Cousineau (Cousineau, 2005). *p ≤ .05, ***p ≤ .001.

We were also interested in any potential differences in the presence of the biomechanical indexes between CRPS patients and control participants, and more specifically whether the MOLA effect could be modulated by the group and/or laterality factors. The ANOVA indeed revealed that *group* and *rotation* interacted with the *orientation* factor (F(2.33,70.06) = 3.51, *p* = .029, η^2^_P_ = .1). *Group* and *rotation* furthermore interacted with the *laterality* factor (F(1,30) = 5.838, *p* = .022, η^2^_P_ = .163). However, since these factors did not interact with *stimulus type*, these effects did not dissociate hand stimuli from non-bodily stimuli (see Table S2). Contrast analyses can be visualized in Fig. 4 and are detailed in the Supplementary material.

**Figure 4.**
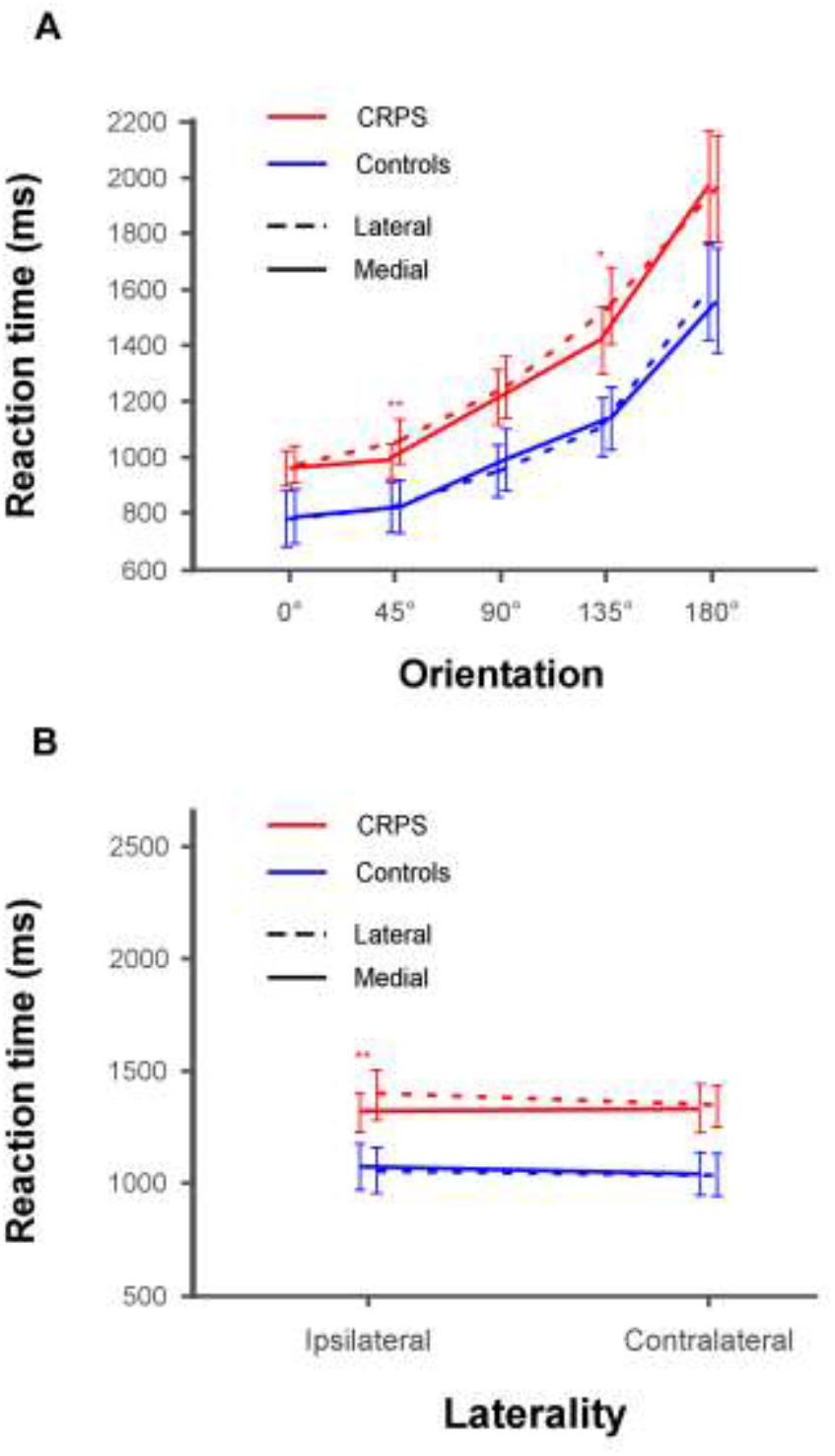
Mean RTs according to the group, the rotation and the orientation or laterality of the stimulus. **A**. Illustration of the interaction between group, rotation and orientation. The lines represent the comparison of RTs between the lateral (dashed lines) and medial (plain lines) rotations for both CRPS (red) and control (blue) participants as a function of the orientation of the stimuli (i.e. 0° vs. 45° vs. 90° vs. 135° vs. 180°). **B**. Illustration of the interaction between group, rotation and laterality. The lines represent the comparison of RTs between the lateral (dashed lines) and medial (plain lines) rotations for both CRPS (red) and control (blue) participants as a function of the laterality of the stimuli (i.e. ipsilateral vs. contralateral). Error bars represent the 95% confidence intervals adapted according to the method of Cousineau (Cousineau, 2005). *p ≤ .05, **p ≤ .01.

Finally, we also tested whether the task instruction could modulate RTs, specifically for hand stimuli, since matching judgements, as compared to laterality, i.e. spatial, judgments, are thought to rather rely on visual imagery processes. The results of the ANOVA showed an interaction between *stimulus type* and *task* (F(1.48,44.48) = 4.50, *p* = .026, η^2^_P_ = .13), indicating that for hand stimuli, RTs were significantly slower for the laterality judgement task than for the matching task (F(1,30) =5.37, *p* = .028, η^2^_P_ = .15). Such a difference was not significant for the house and letter stimuli (all F ≤ .05, *p* ≥ .828, η^2^_P_ ≤ .01).

### 3.2 Experiment 2: Apparent body movement perception task

The ANOVA revealed a main effect of the *group* (F(1,23) = 4.61, *p* = .043, η^2^_P_ = .17), which also significantly interacted with *speed* (F(4,92) = 5.63, *p* < .001, η^2^_P_ = .20; Fig. 6). Contrast analyses showed that while in the control group the performance was significantly influenced by *speed* (F(2.23,26.82) = 8.85, *p* = .001, η^2^_P_ = .42), this was not the case in the CRPS group (F(2.17,23.89) = .74, *p* = .497, η^2^_P_ = .06). In the control group, the percentage of correct paths increased progressively and significantly from speed 1 to 3 and plateaued from speed 3 to 5 (speed 1 vs. speed 2: F(1,12) = 10.89, *p* = .006, η^2^_P_ = .48; speed 2 vs. speed 3: F(1,12) = 5.79, *p* = .033, η^2^_P_ = .33; no significant difference for the comparisons from 3 to 5: all F ≤ 1.73, *p* ≥ .212, η^2^_P_ ≤ .13).

Importantly, the percentage of long paths was significantly lower in the CRPS than in the control participants for all speed (speed 2: F(1,23) = 4.86, *p* = .038, η^2^_P_ = .17; speed 3: F(1,23) = 5.59, *p* = .027, η^2^_P_ = .20; speed 4: F(1,23) = 4.59, *p* = .043, η^2^_P_ = .17; speed 5: F(1,23) = 7.95, *p* = .010, η^2^_P_ = .26), with the exception of speed 1 (F(1,23) = .042, *p* = .839, η^2^_P_ = .00) (Fig. 5).

**Figure 5.**
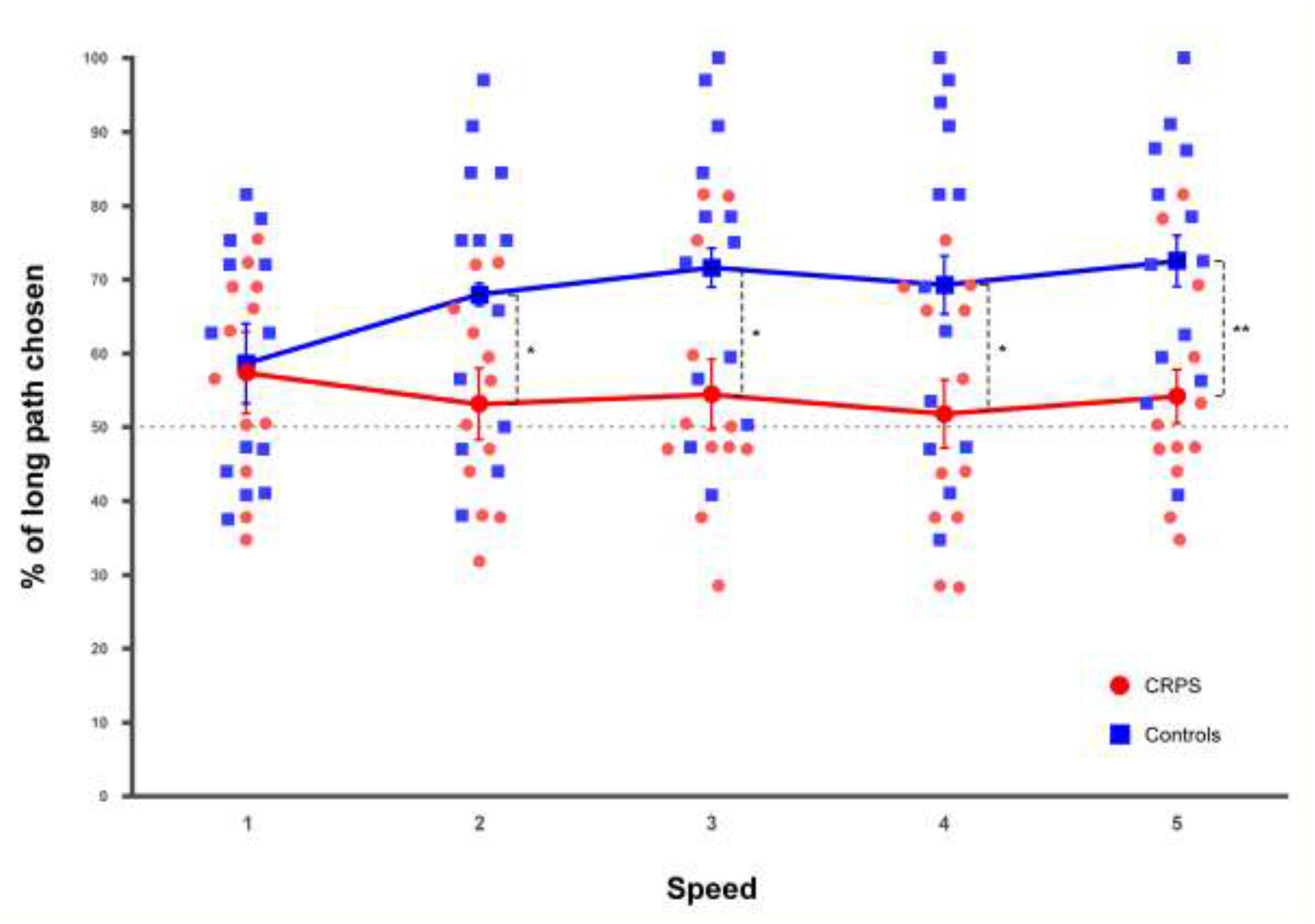
Mean percentage of the long path chosen for the different speed (1 to 5). CRPS patients are represented in red circles and control participants in blue squares. Smaller and lighter colored circles/squares represent the individual data. Error bars indicate the 95% confidence intervals adapted according to the method of Cousineau (Cousineau, 2005). Asterisks in squares indicate t-tests significantly different from 50. *p ≤ .05, **p ≤ .01.

However, no modulation effect of the laterality factor could be evidenced, since none of the other main effects or interactions reached significance (all F ≤ 2.09, *p* ≥ .089, η^2^_P_ ≤ .08).

The paired-samples t-tests completed this finding, by showing that for none of the speed the CRPS patients were more likely to perceive the longer path than the shorter path (all t(11) ≤ 1.85, *p* ≥ .092, d ≤ .53), whereas in the control group the participants chose the long path significantly more often than the short path for all speed (all t(11) > 3.02, *p* ≤ .011, d ≤ .84), with the exception of speed 1 (t(11) = 1.92, *p* = .079, d = .53) (Fig. 5).

## 4. Discussion

In the present study we investigated the abilities of CRPS patients to represent and perceive their affected limb, by specifically focusing on the representation of the biomechanical constraints of the upper limbs and their impact on perceptual judgments. To this aim, CRPS and matched controls performed both a hand laterality judgment (HLT) and apparent body movement perception task. We hypothesized that CRPS patients, as compared to controls, would have difficulties in judging stimuli corresponding to their affected limb. Previous studies have regularly measured body representation with the HLT in CRPS and have indeed shown deficits for stimuli corresponding to the affected limb, which has been interpreted as distortions in hand schema representation (e.g. Moseley, 2004b; Reid *et al*., 2016). These unilateral cognitive distortions have been hypothesized to be linked to findings of lateralized structural and functional changes in the cortical sensorimotor representations of the affected limb (Schwenkreis *et al*., 2009; Birklein and Dimova, 2017).

In the first experiment with the HLT we indeed showed that CRPS patients were significantly slower than controls in judging the laterality of hand stimuli, whereas there was no difference for the control stimuli, i.e. houses and letters. Importantly, and contrary to our hypotheses, this effect was not lateralized, i.e. patients were slower than controls irrespective of whether they judged hand stimuli corresponding to their affected or unaffected hand.

At first glance, this finding seems to contradict the existing literature, especially in light of the unilateral cortical changes in primary sensory and motor cortices that have been described in CRPS. However, even though performing implicit motor imagery on hands during the HLT recruits an extensive cortical network that includes frontal motor and premotor structures (with the contribution of the primary motor cortex being less clear, see e.g. de Lange *et al*., 2005; Bode *et al*., 2007), as well as the basal ganglia, particularly the posterior parietal cortex seems crucially involved (Bonda *et al*., 1995; Parsons *et al*., 1995; Kosslyn *et al*., 1998; de Lange *et al*., 2005). Indeed, both actual and imagined movements are thought to depend on the body schema, with the posterior parietal cortex considered as an integral component of the neural basis underlying body schema representation (Schwoebel and Coslett, 2005). Yet, there is not much consistent evidence for parietal changes in CRPS, besides from what has been inferred from behavioral results. Whereas some studies showed for example weaker posterior parietal activations in response to tactile stimulation applied on the affected and unaffected hand in CRPS as compared to controls (Vartiainen *et al*., 2008; Kuttikat *et al*., 2018), others showed greater bilateral activations than controls in the intraparietal sulcus during finger movement of the affected hand, which correlated with the degree of motor impairment (Maihofner *et al*., 2007). Reduced grey matter volume in the right inferior parietal lobule in early-stage (but not late-stage) CRPS as compared to healthy participants has also been described (Shokouhi *et al*., 2018). Interestingly, a recent study (Kohler *et al*., 2019) did not show any difference between healthy controls and CRPS patients in the activation of the typical fronto-parietal network during a HLT. Instead, CRPS patients showed reduced activity in subcortical areas, such as the subthalamic nucleus, nucleus accumbens and bilateral putamen. Importantly, in this same study, the RTs of CRPS patients were significantly slower than those of the healthy controls, but without any lateralization effect for the side of the affected limb.

Such non-lateralized slowness of CRPS patients in the HLT is also corroborated by other behavioral studies (Reinersmann *et al*., 2010; Bultitude *et al*., 2017; Wittayer *et al*., 2018). These findings have been hypothesized to possibly reflect non-specific sustained attention difficulties in CRPS rather than deficits in body representation. Our results however show that this explanation is very unlikely, since the slowed hand judgments of patients with regard to the controls did not generalize to letter and house stimuli judgments. It is interesting to note that there are also studies using body representation tasks other than the HLT which indicate that deficits in CRPS might extend to the representation and use of the unaffected limb (e.g. Lewis *et al*., 2010; Brun *et al*., 2019).

The ability of the HLT to measure the integrity of the body schema relies on the activation of motor imagery processes during the task, which are generally hypothesized to be highlighted by the presence of biomechanical indexes in the RT. However, a switch to non-motor related strategies to solve the task is not uncommon, which could potentially explain inconsistencies between CRPS studies that employed the HLT task. Our study therefore also focused on analyzing the presence of typical biomechanical indexes in the RT data, specifically with the MOLA effect (TR medial ≠ TR lateral). We indeed found some interactions with the rotation factor, which seemed however rather due to minor differences and not particularly relevant for our questions. The MOLA effect should generally only be observed for hand stimuli, since letter and house stimuli are not supposed to trigger motor imagery. Yet, the MOLA effect was only in very specific conditions observed for hand stimuli and, surprisingly, also for house stimuli. Globally there were no systematic differences between CRPS patients and controls. Only the results regarding task instruction suggested that both CRPS and controls might have used a motor imagery for the hand stimuli when they performed the laterality task. Indeed, RT were significantly different between the laterality judgment and the matching task for hand stimuli, suggesting that different mental imagery strategies might have been used, namely motor and visual imagery (Hoyek *et al*., 2014), respectively.

Based on these inconclusive results we are not able to confirm that our HLT measured motor imagery performance of the participants and that differences between CRPS and controls are explained by body schema representation. The possibility that patients and controls used alternative strategies to perform the task can indeed not be excluded. There are different possible reasons why participants might have used an alternative strategy. Patients could have for example used a visual imagery strategy to compensate for their difficulties in hand schema representation or to avoid an increase in pain (see e.g. King *et al*., 2015). However, this does not explain why controls did also not show the typical indexes of biomechanical constraints. There are also several experimental factors that might have made it difficult to prime motor imagery with the task. What could be potentially problematic is the use of only one hand view. One could hypothesize that, in this case, participants easily become experts in the task or use other strategies, so that it would not be necessary for them anymore to mentally manipulate the hands to judge their laterality. It has indeed been shown that the MOLA effect cannot always be evidenced when hand stimuli are presented from only one point of view (i.e. the back view, ter Horst *et al*., 2010). Also, although the order of the different blocks was randomized, the fact that some participants started the task with house or letter judgments, priming the use of visual imagery, could have influenced the strategy for subsequent hand judgements. A recent study furthermore showed that a motor imagery strategy is not universally and specifically used to perform the HLT (Mibu *et al*., 2020).

Another critical point that has been raised in recent years is that the presence of the indexes of biomechanical constraints might actually not automatically reflect the activation of motor imagery processes during the task. There are indeed studies that showed that the effect of biomechanical constraints on laterality judgments can also be observed in motor-impaired individuals (e.g. Fiorio *et al*., 2005) and individuals with a congenital absence of upper limbs (Vannuscorps *et al*., 2012; Vannuscorps and Caramazza, 2015). That we observed the MOLA effect for house stimuli also supports this hypothesis. These different arguments show that it might be challenging to test motor imagery and the integrity of biomechanical constraint representation with the HLT.

Participants therefore also performed a second task which specifically tests the representation of the biomechanical constraints of the upper limbs and their influence on perceptual judgments. This apparent body movement perception task is based on the apparent motion paradigm (Shiffrar and Freyd, 1990; Chatterjee *et al*., 1996), which postulates that if participants observe two sequentially presented objects, they typically report seeing motion along the shortest possible path between the two objects. If however pictures of an actor whose hand alternates between two positions are shown, the perception follows biomechanically plausible paths rather than a path along the shortest distance, in particular if the shorter path between the two hand positions is biomechanically impossible. The implicit knowledge and representation of the biomechanical constraints is thus supposed to influence the perception of the actor’s movements, corresponding to a biomechanical bias in apparent motion perception (Vannuscorps and Caramazza, 2016). In the present study the shorter path always corresponded to a biomechanically implausible movement, whereas the longer path was always biomechanically plausible. If the participants use a correct knowledge of their biomechanical constraints for their perceptual judgments, the long path should be chosen most of the time. Our results showed that the control participants chose the long, i.e. biomechanically plausible, path significantly more often than did the CRPS patients, with an exception for the fastest speed of presentation. The literature indeed shows that the frequency of choosing the plausible path is modulated by stimulus exposure duration and ISI, i.e. the interval between the two pictures needs to be long enough for the longer movement to be perceived as plausible (Vannuscorps and Caramazza, 2016). Importantly, CRPS patients had systematically equal chances to choose the biomechanically plausible or the implausible path, independently of the presentation speed/ISI and of the laterality of the depicted upper limbs.

These results suggest that CRPS patients might have difficulties in representing or perceiving the biomechanics of the upper limbs. Whether this is due to CRPS-related changes in cortical sensorimotor areas or more generalized changes in body representation needs to be determined. Vannuscorps and Caramazza (2016) for example demonstrated that even participants born without upper limbs can show the typical indexes of the influence of the biomechanical bias in apparent motion perception, suggesting that rather than relying on online sensorimotor representations and motor simulation, the performance on the task might depend on a visual perceptual or semantic knowledge of how a body should move.

Considering the results of the two experiments, the data does thus not allow to conclude on deficits in motor imagery abilities in CRPS. We clearly showed that patients process hand stimuli differently than controls in the HLT, but since for both patients and controls the typical indexes of biomechanical constraints were not observed and slowed responses in patients were not lateralized, there is no reason to believe that it would be specifically the online sensorimotor representation of the affected limb that is disturbed in CRPS. This is also corroborated by the observation that patients are slower than controls in judging hand stimuli independently of the task, i.e. even in the matching task that is supposed to induce visual imagery instead of motor imagery. The results of the apparent body movement perception task also support this hypothesis, showing that, although the perception and/or use of the biomechanics seems to be disturbed in CRPS, patients do not systematically process hands as any other object, since they perceive the shorter and the longer path equally often. This does of course not exclude the possibility of difficulties related to body schema representation in CRPS, but shows that by using the HLT, in our and other studies, there is a certain uncertainty about what is exactly assessed.

This is particularly relevant considering that laterality judgments have been used in the rehabilitation of CRPS and other chronic pain conditions, notably with the aim to activate and restore motor imagery and the cortical (motor) representation of the limb, which would in turn improve pain and other CRPS-related symptoms (e.g. Moseley, 2004a; Moseley and Flor, 2012). Yet, the ability of these tasks to elicit motor imagery is critical to their use in therapy. If this is not the case, we might simply train patients to treat hands as simple objects. Our results show the necessity of questioning what and how exactly we are rehabilitating with these tasks, also in the context of understanding why motor imagery based programs might not always be effective in decreasing pain (e.g. Johnson *et al*., 2012).

To conclude, our results indicate that upper-limb CRPS patients can present with non-lateralized impairments of hand representation, which might be partly related to difficulties in representing and perceiving the biomechanics of their upper limbs. At the same time, and most importantly, the present results also highlight the difficulty of reliably measuring motor imagery and the importance of verifying the mechanisms and strategies that underlie the patients’ performance on the HLT, which should lead to more cautious interpretations, especially in a rehabilitative perspective.

## Data Availability

The datasets generated during and/or analysed during the current study are available from
the corresponding author on reasonable request.

## Acknowledgements

L.F., C.V. and V.L. are supported by the Fund for Scientific Research of the French-speaking Community of Belgium (F.R.S.-FNRS). We thank the staffs of the Departments of Physical Medicine and of Orthopedics of the Saint-Luc Hospital (Brussels), of the Rehabilitation Center of the Erasme Hospital (Brussels), and of the Department of Physical Medicine of the Bois de la Pierre Clinical Center (Wavre) for their help in recruiting the participants.

## References

Birklein F, Dimova V. Complex regional pain syndrome-up-to-date. Pain Rep 2017; 2(6): e624–e.

Birklein F, Schlereth T. Complex regional pain syndrome-significant progress in understanding. Pain 2015; 156 Suppl 1: S94–103.

Bode S, Koeneke S, Jäncke L. Different strategies do not moderate primary motor cortex involvement in mental rotation: a TMS study. Behavioral and Brain Functions 2007; 3(1): 38.

Bonda E, Petrides M, Frey S, Evans A. Neural correlates of mental transformations of the body-in-space. Proceedings of the National Academy of Sciences 1995; 92(24): 11180–4.

Breimhorst M, Dellen C, Wittayer M, Rebhorn C, Drummond PD, Birklein F. Mental load during cognitive performance in complex regional pain syndrome I. Eur J Pain 2018; 22(7): 1343–50.

Brugger P, Brugger S. The Easter Bunny in October: Is it Disguised as a Duck? 1993; 76(2): 577–8.

Brun C, Giorgi N, Pinard AM, Gagne M, McCabe CS, Mercier C. Exploring the Relationships Between Altered Body Perception, Limb Position Sense, and Limb Movement Sense in Complex Regional Pain Syndrome. The journal of pain 2019; 20(1): 17–27.

Bultitude JH, Walker I, Spence C. Biased covert visual attention to body and space in complex regional pain syndrome. Brain 2017; 140(9): 2306–21.

Chatterjee HS, Freyd JJ, Shiffrar M. Configural processing in the perception of apparent biological motion. Journal of Experimental Psychology: Human Perception and Performance 1996; 22(4): 916–29.

Cooper LA, Shepard RN. Mental transformation in the identification of left and right hands. Journal of Experimental Psychology: Human Perception and Performance 1975; 1(1): 48–56.

Cousineau D. Confidence intervals in within-subject designs: a simpler solution to Loftus and Masson’s method. Tutor Quant Methods Psychol 2005; 1(1): 42–5.

Daprati E, Nico D, Duval S, Lacquaniti F. Different motor imagery modes following brain damage. Cortex 2010; 46(8): 1016–30.

de Lange FP, Hagoort P, Toni I. Neural Topography and Content of Movement Representations. Journal of Cognitive Neuroscience 2005; 17(1): 97–112.

Fiorio M, Tinazzi M, Aglioti SM. Selective impairment of hand mental rotation in patients with focal hand dystonia. Brain 2005; 129(1): 47–54.

Funk M, Brugger P. Mental rotation of congenitally absent hands. Journal of the International Neuropsychological Society 2008; 14(1): 81–9.

Halicka M, Vittersø AD, Proulx MJ, Bultitude JH. Neuropsychological Changes in Complex Regional Pain Syndrome (CRPS). Behavioural Neurology 2020; 2020: 1–30.

Harden RN, Bruehl S, Perez RS, Birklein F, Marinus J, Maihofner C, et al. Validation of proposed diagnostic criteria (the “Budapest Criteria”) for Complex Regional Pain Syndrome. Pain 2010; 150(2): 268–74.

Hoyek N, Di Rienzo F, Collet C, Creveaux T, Guillot A. Hand mental rotation is not systematically altered by actual body position: Laterality judgment versus same–different comparison tasks. Attention, Perception, & Psychophysics 2014; 76(2): 519–26.

Ionta S, Fourkas AD, Fiorio M, Aglioti SM. The influence of hands posture on mental rotation of hands and feet. Exp Brain Res 2007; 183(1): 1–7.

Johnson S, Hall J, Barnett S, Draper M, Derbyshire G, Haynes L, et al. Using graded motor imagery for complex regional pain syndrome in clinical practice: Failure to improve pain. European Journal of Pain 2012; 16(4): 550–61.

Juottonen K, Gockel M, Silén T, Hurri H, Hari R, Forss N. Altered central sensorimotor processing in patients with complex regional pain syndrome. Pain 2002; 98: 315–23.

King R, Johnson MI, Ryan CG, Robinson V, Martin DJ, Punt TD. My Foot? Motor Imagery-Evoked Pain, Alternative Strategies and Implications for Laterality Recognition Tasks. Pain Medicine 2015; 16(3): 555–7.

Kohler M, Strauss S, Horn U, Langner I, Usichenko T, Neumann N, et al. Differences in Neuronal Representation of Mental Rotation in Patients With Complex Regional Pain Syndrome and Healthy Controls. The Journal of Pain 2019; 20(8): 898–907.

Kosslyn SM, Digirolamo GJ, Thompson WL, Alpert NM. Mental rotation of objects versus hands: Neural mechanisms revealed by positron emission tomography. Psychophysiology 1998; 35(2): 151–61.

Krause P, Forderreuther S, Straube A. TMS motor cortical brain mapping in patients with complex regional pain syndrome type I. Clinical neurophysiology 2006; 117(1): 169–76.

Kuttikat A, Noreika V, Chennu S, Shenker N, Bekinschtein T, Brown CA. Altered Neurocognitive Processing of Tactile Stimuli in Patients with Complex Regional Pain Syndrome. The journal of pain 2018; 19(4): 395–409.

Lewis JS, Kersten P, McPherson KM, Taylor GJ, Harris N, McCabe CS, et al. Wherever is my arm? Impaired upper limb position accuracy in complex regional pain syndrome. Pain 2010; 149(3): 463–9.

Maihofner C, Baron R, DeCol R, Binder A, Birklein F, Deuschl G, et al. The motor system shows adaptive changes in complex regional pain syndrome. Brain 2007; 130(Pt 10): 2671-87.

Maihofner C, Handwerker HO, Neundorfer B, Birklein F. Patterns of cortical reorganization in complex regional pain syndrome. Neurology 2003; 61(12): 1707–15.

Maihofner C, Handwerker HO, Neundorfer B, Birklein F. Cortical reorganization during recovery from complex regional pain syndrome. Neurology 2004; 63(4): 693–701.

Mancini F, Wang AP, Schira MM, Isherwood ZJ, McAuley JH, Iannetti GD, et al. Fine-Grained Mapping of Cortical Somatotopies in Chronic Complex Regional Pain Syndrome. J Neurosci 2019; 39(46): 9185–96.

Marinus J, Moseley GL, Birklein F, Baron R, Maihöfner C, Kingery WS, et al. Clinical features and pathophysiology of complex regional pain syndrome. Lancet Neurol 2011; 10(7): 637–48.

Mibu A, Kan S, Nishigami T, Fujino Y, Shibata M. Performing the hand laterality judgement task does not necessarily require motor imagery. Scientific reports 2020; 10(1): 5155.

Moseley GL. Graded motor imagery is effective for long-standing complex regional pain syndrome: a randomised controlled trial. Pain 2004a; 108(1-2): 192–8.

Moseley GL. Why do people with complex regional pain syndrome take longer to recognize their affected hand? Neurology 2004b; 62: 2182–6.

Moseley GL, Butler DS, Beames TB, Giles TJ. The graded motor imagery handbook. Adelaide, Australia: Noigroup Publications; 2012.

Moseley GL, Flor H. Targeting cortical representations in the treatment of chronic pain: a review. Neurorehabil Neural Repair 2012; 26(6): 646–52.

Nordin M, Frankel VH. Basic biomechanics of the musculoskeletal system, 3rd ed. Philadelphia: Lippincott Williams and Wilkins; 2001.

Parsons LM. Imagined spatial transformations of one’s hands and feet. Cognitive Psychology 1987; 19(2): 178–241.

Parsons LM. Temporal and kinematic properties of motor behavior reflected in mentally simulated action. Journal of Experimental Psychology: Human Perception and Performance 1994; 20(4): 709–30.

Parsons LM, Fox PT, Downs JH, Glass T, Hirsch TB, Martin CC, et al. Use of implicit motor imagery for visual shape discrimination as revealed by PET. Nature 1995; 375(6526): 54–8.

Pfannmoller J, Strauss S, Langner I, Usichenko T, Lotze M. Investigations on maladaptive plasticity in the sensorimotor cortex of unilateral upper limb CRPS I patients. Restor Neurol Neurosci 2019; 37(2): 143–53.

Pleger B, Draganski B, Schwenkreis P, Lenz M, Nicolas V, Maier C, et al. Complex regional pain syndrome type I affects brain structure in prefrontal and motor cortex. PLoS One 2014; 9(1): e85372.

Pleger B, Ragert P, Schwenkreis P, Forster AF, Wilimzig C, Dinse H, et al. Patterns of cortical reorganization parallel impaired tactile discrimination and pain intensity in complex regional pain syndrome. NeuroImage 2006; 32(2): 503–10.

Pleger B, Tegenthoff M, Ragert P, Forster AF, Dinse HR, Schwenkreis P, et al. Sensorimotor retuning [corrected] in complex regional pain syndrome parallels pain reduction. Annals of neurology 2005; 57(3): 425–9.

Pleger B, Tegenthoff M, Schwenkreis P, Janssen F, Ragert P, Dinse HR, et al. Mean sustained pain levels are linked to hemispherical side-to-side differences of primary somatosensory cortex in the complex regional pain syndrome I. Exp Brain Res 2004; 155(1): 115–9.

Reid E, Wallwork SB, Harvie D, Chalmers KJ, Gallace A, Spence C, et al. A New Kind of Spatial Inattention Associated With Chronic Limb Pain? Annals of neurology 2016; 79(4): 701–4.

Reinersmann A, Haarmeyer GS, Blankenburg M, Frettloh J, Krumova EK, Ocklenburg S, et al. Left is where the L is right. Significantly delayed reaction time in limb laterality recognition in both CRPS and phantom limb pain patients. Neuroscience letters 2010; 486(3): 240–5.

Reinersmann A, Landwehrt J, Krumova EK, Ocklenburg S, Gunturkun O, Maier C. Impaired spatial body representation in complex regional pain syndrome type 1 (CRPS I). Pain 2012; 153(11): 2174–81.

Reinersmann A, Maier C, Schwenkreis P, Lenz M. Complex regional pain syndrome: more than a peripheral disease. Pain Management 2013; 3(6): 495–502.

Schwenkreis P, Maier C, Tegenthoff M. Functional imaging of central nervous system involvement in complex regional pain syndrome. AJNR American journal of neuroradiology 2009; 30(7): 1279–84.

Schwoebel J, Buxbaum LJ, Branch Coslett H. Representations of the human body in the production and imitation of complex movements. Cognitive Neuropsychology 2004; 21(2-4): 285-98.

Schwoebel J, Coslett HB. Evidence for Multiple, Distinct Representations of the Human Body. Journal of Cognitive Neuroscience 2005; 17(4): 543–53.

Schwoebel J, Coslett HB, Bradt J, Friedman R, Dileo C. Pain and the body schema: Effects of pain severity on mental representations of movement. Neurology 2002; 59(5): 775–7.

Schwoebel J, Friedman R, Duda N, Coslett HB. Pain and the body schema. Evidence for peripheral effects on mental representations of movement. Brain 2001; 124: 2098–104.

Shepard RN, Metzler J. Mental Rotation of Three-Dimensional Objects. Science 1971; 171(3972): 701–3.

Shiffrar M, Freyd JJ. Apparent Motion of the Human Body. Psychological Science 1990; 1(4): 257–64.

Shokouhi M, Clarke C, Morley-Forster P, Moulin DE, Davis KD, St Lawrence K. Structural and Functional Brain Changes at Early and Late Stages of Complex Regional Pain Syndrome. The journal of pain 2018; 19(2): 146–57.

Sirigu A, Duhamel JR. Motor and Visual Imagery as Two Complementary but Neurally Dissociable Mental Processes. J Cogn Neurosci 2001; 13(7): 910–9.

ter Horst AC, van Lier R, Steenbergen B. Mental rotation task of hands: differential influence number of rotational axes. Experimental Brain Research 2010; 203(2): 347–54.

Vannuscorps G, Caramazza A. Typical biomechanical bias in the perception of congenitally absent hands. Cortex 2015; 67: 147–50.

Vannuscorps G, Caramazza A. The origin of the biomechanical bias in apparent body movement perception. Neuropsychologia 2016; 89: 281–6.

Vannuscorps G, Pillon A, Andres M. Effect of biomechanical constraints in the hand laterality judgment task: where does it come from? Front Hum Neurosci 2012; 6(299).

Vartiainen NV, Kirveskari E, Forss N. Central processing of tactile and nociceptive stimuli in complex regional pain syndrome. Clinical neurophysiology 2008; 119(10): 2380–8.

Wittayer M, Dimova V, Birklein F, Schlereth T. Correlates and importance of neglect-like symptoms in complex regional pain syndrome. Pain 2018; 159(5): 978–86.

