## Supplementary material for "Atypical influence of biomechanical knowledge in Complex Regional Pain Syndrome-Towards a different perspective on body representation"

Supplementary materials

### 1. Pain ratings

#### 1.1 Measures and data analyses

**1.1.1** To test whether the subjective pain ratings were influenced by the stimulus type and/or the task, we computed the average of the post - pre block pain ratings difference for each stimulus type x task combination in the CRPS group. Pain ratings were scored from 0 (no pain) to 10 (worst pain imaginable). A non-parametric Friedman test on the data of the CRPS group with the combination of stimulus type x task (Hand + laterality judgment vs. Hand + matching vs. House + laterality judgment vs. House + matching vs. Letter + laterality judgment vs. Letter + matching) as within participant factor was performed.

**1.1.2** Furthermore, to test whether the magnitude of pain during a specific experimental block predicted the CRPS patients' performance on this block, the subjective pain ratings for each patient were averaged through pre and post block pain ratings for each stimulus type x task combination in the CRPS group. Linear regression was performed to test whether average pain could predict CRPS patients' RTs and/or accuracy in each of the possible stimulus type x task combinations.

#### 1.2 Results

**1.2.1** The subjective pain ratings of CRPS patients did not significantly differ between the different stimulus type x task combinations ($\chi^2(5)= 1.47$, $p = .917$).

**1.2.2** RTs for none of the different stimulus type and task combinations were significantly predicted by patients' subject pain ratings (all $r^2 \leq .36$, $F \leq 2.12$, $p \geq .167$), with an exception for the condition presenting house stimuli combined with the matching task which approached significance ($r^2= .47$, $F(1,15)= 3.96$, $p = .066$). Accuracy was generally also not predicted by the subjective pain ratings (all $r^2 \leq .33$, $F \leq 1.72$, $p \geq .210$), with the exception of the condition combining letter stimuli and the matching task ($r^2= .51$, $F(1,15)= 4.86$, $p = .045$).

### 2. Additional results for RTs

#### 2.1 Supplementary contrast analysis for the *group x stimulus type* interaction

In both groups there was a significant difference in RTs between the three types of stimuli (CRPS: $F(1.2,17.97)= 46.32$, $p = .000$, $\eta^2_P = .75$; Controls: $F(1.38,20.74) = 17.91$, $p = .000$, $\eta^2_P = .54$). Indeed, in both groups, hands were judged slower than house (CRPS: $F(1,15)= 55.14$, $p = .000$, $\eta^2_P = .79$; Controls: $F(1,15) = 22.48$, $p = .000$, $\eta^2_P = .60$) and letter stimuli (CRPS: $F(1,15) = 43.31$, $p = .000$, $\eta^2_P = .74$; Controls: $F(1,15) = 14.05$, $p = .002$, $\eta^2_P = .48$). In both groups letter stimuli were also judged slower than house stimuli (CRPS: $F(1,15) = 7.89$, $p = .013$, $\eta^2_P = .34$; Controls: $F(1,15) = 10.38$, $p = .006$, $\eta^2_P = .41$).

#### 2.2 Supplementary contrast analysis for the *group x rotation x orientation* interaction

Contrast analyses showed that for stimuli turned at 45° and 135°, medial rotations were judged faster than lateral rotations in the CRPS group (45°: $F(1,15) = 9.40$, $p = .008$, $\eta^2_P = .38$; 135°: $F(1,15) = 5.73$, $p = .03$, $\eta^2_P = .28$). There was neither a rotation effect for these orientations in the control group, nor for any of the other orientations in any of the two groups (all $F \leq 4.09$, $p \geq .061$, $\eta^2_P \leq .21$).

#### 2.3 Supplementary contrast analysis for the *group x rotation x laterality* interaction

Contrast analyses showed that for ipsilateral stimuli, the CRPS group was faster in judging medially than laterally rotated stimuli ($F(1,15) = 10.24$, $p = .006$, $\eta^2_P = .41$). Such a significant rotation effect was neither observed for the same ipsilateral stimuli in the control group, nor for the contralateral stimuli in any of the two groups (all $F \leq 1.15$, $p \geq .300$, $\eta^2_P \leq .07$).

### 3. Results for the accuracy

The ANOVA revealed a significant interaction between the *group* and *laterality* ($F(1,30) = 6.38$, $p = .017$, $\eta^2_P = .17$). Contrasts analyses showed that CRPS patients were significantly less accurate (M = 93.7± 4.2%) than the control participants (M = 97.3±1.5%) in judging ipsilateral stimuli ($F(1,30) = 6.18$, $p = .019$, $\eta^2_P = .17$), that is, pictures of hands corresponding to their affected limb and images of houses and letters with target details in the side corresponding to their affected limb. There was no significant difference between the groups for contralateral stimuli ($F(1,30) = 1.89$, $p = .180$, $\eta^2_P = .06$). CRPS patients were in general more accurate for contralateral (M = 95.1±3.9%) than for ipsilateral stimuli (M = 93.7±4.2%; $F(1,15) = 9.22$, $p = .008$, $\eta^2_P = .38$), while such a difference was not significant in the control group ($F(1,15) = .36$, $p = .555$, $\eta^2_P = .02$).

Analyses of accuracy also revealed a significant main effect of *stimulus type* ($F(2,60) = 7.15$, $p = .002$, $\eta^2_P = .19$), suggesting that all participants were significantly more accurate in performing the task with images of houses (M = 98.1±1.5%) than with pictures of hands (M = 93.5% ± 4.9%; $F(1,30) = 16.19$, $p = .000$, $\eta^2_P = .35$) and images of letters (M = 95.8±4.6%; $F(1,30) = 4.78$, $p = .037$, $\eta^2_P = .14$). There was no significant difference in accuracy between hand and letter stimuli ($F(1,30) = 2.59$, $p = .118$, $\eta^2_P = .08$). There was furthermore a significant main effect of *orientation* ($F(1.07,32.00) = 19.41$, $p < .001$, $\eta^2_P = .39$), showing that the participants' performance significantly decreased throughout 45° (M = 98.6±1.3%), 90° (M = 97.9±1.6%), 135° (M = 96.4±2.4%) and 180° (M = 87.7±10%) orientations (all paired comparisons: $F \geq 10.37$, $p \leq .003$, $\eta^2_P \geq .26$; except between 0° (M = 98.4±1.3%) and 45° orientations: $F(1,30) = 1.31$, $p = .261$, $\eta^2_P = .04$).

### 4. Supplementary tables

Supplementary Table S1

Results of the ANOVA on the accuracy data with *stimulus type* (hand vs. house vs. letter), *laterality* (*ipsilateral vs. contralateral*), *rotation* (medial vs. lateral) and *orientation* (0° vs. 45° vs. 90° vs. 135° vs. 180°) as within-participants factors and *group* (laterality judgment vs. matching) as between-participant factor.

| **Factors** | **F** | ***p*** | **$\eta^2_p$** |
|---|---|---|---|
| *Group* | 3.99 | .055 | .12 |
| *Stimulus* | 7.15 | .002 | .19 |
| *Stimulus × Group* | 2.29 | .110 | .07 |
| *Task* | 1.79 | .191 | .06 |
| *Task × Group* | .04 | .843 | .00 |
| *Laterality* | 2.71 | .110 | .08 |
| *Laterality × Group* | 6.38 | .017 | .17 |
| *Rotation* | .15 | .703 | .01 |
| *Rotation × Group* | 1.60 | .183 | .06 |
| *Orientation* | 19.41 | .000 | .39 |

| | | | |
|---|---|---|---|
| *Orientation* × *Group* | 2.71 | .107 | .08 |
| *Stimulus* × *Task* | 2.52 | .089 | .08 |
| *Stimulus* × *Task* × *Group* | .71 | .495 | .02 |
| *Stimulus* × *Laterality* | .41 | .601 | .01 |
| *Stimulus* × *Laterality* × *Group* | 1.47 | .239 | .05 |
| *Stimulus* × *Rotation* | 1.43 | .247 | .05 |
| *Stimulus* × *Rotation* × *Group* | .16 | .789 | .00 |
| *Stimulus* × *Orientation* | 1.15 | .316 | .04 |
| *Stimulus* × *Orientation* × *Group* | .82 | .424 | .03 |
| *Task* × *Laterality* | .01 | .906 | .000 |
| *Task* × *Laterality* × *Group* | 1.72 | .200 | .05 |
| *Task* × *Rotation* | .26 | .612 | .01 |
| *Task* × *Rotation* × *Group* | .00 | .975 | .00 |
| *Task* × *Orientation* | .70 | .455 | .02 |
| *Task* × *Orientation* × *Group* | .12 | .816 | .00 |
| *Stimulus* × *Task* × *Laterality* | .30 | .684 | .01 |

| | | | |
|---|---|---|---|
| *Stimulus × Task × Laterality × Group* | .09 | .866 | .00 |
| *Stimulus × Task × Rotation* | 1.54 | .226 | .05 |
| *Stimulus × Task × Rotation × Group* | .03 | .936 | .00 |
| *Stimulus × Task × Orientation* | 2.04 | .125 | .06 |
| *Stimulus × Task × Orientation × Group* | 1.57 | .209 | .05 |
| *Stimulus × Laterality × Rotation* | .78 | .463 | .02 |
| *Stimulus × Laterality × Rotation × Group* | 1.68 | .194 | .05 |
| *Laterality × Rotation* | 1.91 | .178 | .06 |
| *Laterality × Rotation × Group* | .30 | .585 | .01 |
| *Laterality × Orientation* | .60 | .546 | .02 |
| *Laterality × Orientation × Group* | .54 | .574 | .02 |
| *Rotation × Orientation* | .49 | .651 | .02 |
| *Rotation × Orientation × Group* | 2.10 | .119 | .06 |
| *Stimulus × Laterality × Orientation* | .82 | .517 | .03 |
| *Stimulus × Laterality × Orientation × Group* | .52 | .727 | .02 |
| *Stimulus × Rotation × Orientation* | 1.03 | .399 | .03 |

| | | | |
|---|---|---|---|
| *Stimulus × Rotation × Orientation × Group* | 1.32 | .264 | .04 |
| *Task × Laterality × Rotation* | .52 | .476 | .02 |
| *Task × Laterality × Rotation × Group* | 1.24 | .274 | .04 |
| *Task × Laterality × Orientation* | .34 | .755 | .01 |
| *Task × Laterality × Orientation × Group* | 2.42 | .084 | .07 |
| *Task × Rotation × Orientation* | 1.51 | .230 | .05 |
| *Task × Rotation × Orientation × Group* | .24 | .771 | .01 |
| *Laterality × Rotation × Orientation* | 2.72 | .057 | .08 |
| *Laterality × Rotation × Orientation × Group* | 1.44 | .239 | .05 |
| *Stimulus × Task × Laterality × Rotation* | 1.02 | .365 | .03 |
| *Stimulus × Task × Laterality × Rotation × Group* | 1.74 | .185 | .05 |
| *Stimulus × Task × Laterality × Orientation* | .77 | .521 | .02 |
| *Stimulus × Task × Laterality × Orientation × Group* | .40 | .773 | .01 |
| *Stimulus × Task × Rotation × Orientation* | .40 | .801 | .01 |
| *Stimulus × Task × Rotation × Orientation × Group* | .84 | .497 | .03 |
| *Stimulus × Laterality × Rotation × Orientation* | 1.59 | .181 | .05 |

| | | | |
|---|---|---|---|
| *Stimulus × Laterality × Rotation × Orientation × Group* | .93 | .448 | .03 |
| *Task × Laterality × Rotation × Orientation* | .30 | .781 | .01 |
| *Task × Laterality × Rotation × Orientation × Group* | .67 | .543 | .02 |
| *Stimulus × Task × Laterality × Rotation × Orientation* | .28 | .898 | .01 |
| *Stimulus × Task × Laterality × Rotation × Orientation × Group* | 1.36 | .250 | .04 |

Supplementary Table S2

Results of the ANOVA on the RT data with *stimulus type* (hand vs. house vs. letter), *laterality* (*ipsilateral vs. contralateral*), *rotation* (medial vs. lateral) and *orientation* (0° vs. 45° vs. 90° vs. 135° vs. 180°) as within-participants factors and *group* (laterality judgment vs. matching) as between-participant factor.

| Factors | F | *p* | $\eta^2_p$ |
|---|---|---|---|
| *Group* | 7.95 | .008 | .21 |
| *Stimulus* | 63.58 | .000 | .68 |
| *Stimulus* × *Group* | 12.28 | .001 | .29 |
| *Task* | 1.79 | .191 | .06 |
| *Task* × *Group* | 0.07 | .786 | .00 |
| *Laterality* | 2.38 | .134 | .07 |
| *Laterality* × *Group* | 0.17 | .686 | .01 |
| *Rotation* | 3.62 | .067 | .11 |
| *Rotation* × *Group* | 8.38 | .007 | .22 |
| *Orientation* | 197.73 | .000 | .87 |
| *Orientation* × *Group* | 3.21 | .066 | .10 |

| | | | |
|---|---|---|---|
| *Stimulus* × *Task* | 4.49 | .026 | .13 |
| *Stimulus* × *Task* × *Group* | 0.15 | .793 | .00 |
| *Stimulus* × *Laterality* | 2.19 | .131 | .07 |
| *Stimulus* × *Laterality* × *Group* | 0.58 | .530 | .02 |
| *Stimulus* × *Rotation* | 1.25 | .295 | .04 |
| *Stimulus* × *Rotation* × *Group* | 0.37 | .623 | .01 |
| *Stimulus* × *Orientation* | 8.60 | .000 | .22 |
| *Task* × *Laterality* | 0.13 | .725 | .00 |
| *Task* × *Laterality* × *Group* | 2.453 | .128 | .08 |
| *Task* × *Rotation* | 2.08 | .159 | .06 |
| *Task* × *Rotation* × *Group* | 1.89 | .179 | .06 |
| *Task* × *Orientation* | 1.30 | .272 | .04 |
| *Task* × *Orientation* × *Group* | 0.63 | .475 | .02 |
| *Laterality* × *Rotation* | 2.93 | .097 | .09 |
| *Laterality* × *Rotation* × *Group* | 5.84 | .022 | .16 |
| *Laterality* × *Orientation* | 0.40 | .628 | .01 |

| | | | |
|---|---|---|---|
| *Laterality × Orientation × Group* | 0.69 | .477 | .02 |
| *Rotation × Orientation* | 1.21 | .308 | .04 |
| *Rotation × Orientation × Group* | 3.51 | .029 | .10 |
| *Stimulus × Orientation × Group* | 0.61 | .612 | .02 |
| *Stimulus × Task × Laterality* | 0.39 | .678 | .01 |
| *Stimulus × Task × Laterality × Group* | 3.01 | .052 | .09 |
| *Stimulus × Task × Rotation* | 0.75 | .425 | .02 |
| *Stimulus × Task × Rotation × Group* | 2.53 | .112 | .08 |
| *Stimulus × Task × Orientation* | 2.11 | .100 | .07 |
| *Stimulus × Task × Orientation × Group* | 0.63 | .603 | .02 |
| *Stimulus × Laterality × Rotation* | 0.16 | .850 | .00 |
| *Stimulus × Laterality × Rotation × Group* | 1.62 | .206 | .05 |
| *Stimulus × Laterality × Orientation* | 1.16 | .332 | .04 |
| *Stimulus × Laterality × Orientation × Group* | 0.26 | .899 | .01 |
| *Stimulus × Rotation × Orientation* | 3.80 | .004 | .11 |
| *Stimulus × Rotation × Orientation × Group* | 1.29 | .273 | .04 |

| | | | |
|---|---|---|---|
| *Task × Laterality × Rotation* | 1.49 | .232 | .05 |
| *Task × Laterality × Rotation × Group* | 0.02 | .891 | .00 |
| *Task × Laterality × Orientation* | 0.55 | .581 | .02 |
| *Task × Laterality × Orientation × Group* | 0.12 | .887 | .00 |
| *Task × Rotation × Orientation* | 1.66 | .186 | .05 |
| *Task × Rotation × Orientation × Group* | 1.77 | .165 | .06 |
| *Laterality × Rotation × Orientation* | 0.45 | .726 | .01 |
| *Laterality × Rotation × Orientation × Group* | 0.36 | .791 | .01 |
| *Stimulus × Task × Laterality × Rotation* | 0.15 | .798 | .00 |
| *Stimulus × Task × Laterality × Rotation × Group* | 0.05 | .911 | .00 |
| *Stimulus × Task × Laterality × Orientation* | 0.60 | .618 | .02 |
| *Stimulus × Task × Laterality × Orientation × Group* | 0.42 | .740 | .01 |
| *Stimulus × Task × Rotation × Orientation* | 1.54 | .205 | .05 |
| *Stimulus × Task × Rotation × Orientation × Group* | 0.74 | .547 | .02 |
| *Stimulus × Laterality × Rotation × Orientation* | 0.61 | .767 | .02 |
| *Stimulus × Laterality × Rotation × Orientation × Group* | 1.09 | .362 | .03 |

| | | | |
|---|---|---|---|
| *Task × Laterality × Rotation × Orientation* | 0.68 | .545 | .02 |
| *Task × Laterality × Rotation × Orientation × Group* | 0.98 | .420 | .03 |
| *Stimulus × Task × Laterality × Rotation × Orientation* | 1.37 | .245 | .04 |
| *Stimulus × Task × Laterality × Rotation × Orientation × Group* | 0.84 | .208 | .03 |
